# Synthesize Heterogeneous Biological Knowledge via Representation Learning for Alzheimer’s Disease Drug Repurposing

**DOI:** 10.1101/2021.12.03.21267235

**Authors:** Kang-Lin Hsieh, German Plascencia-Villa, Ko-Hong Lin, George Perry, Xiaoqian Jiang, Yejin Kim

## Abstract

Developing drugs for treating Alzheimer’s disease (AD) has been extremely challenging and costly due to limited knowledge on underlying biological mechanisms and therapeutic targets. Repurposing drugs or their combination has shown potential in accelerating drug development due to the reduced drug toxicity while targeting multiple pathologies. To address the challenge in AD drug development, we developed a multi-task deep learning pipeline to integrate a comprehensive knowledge graph on biological/pharmacological interactions and multi-level evidence on drug efficacy, to identify repurposable drugs and their combination candidates. We developed and computationally validated a heterogeneous graph representation model with transfer learning from universal biomedical databases and joint optimization with AD risk genes. Using the drug embedding from the heterogeneous graph representation model, we ranked drug candidates based on evidence from post-treatment transcriptomic patterns, mechanistic efficacy in preclinical models, population-based treatment effect, and Phase II/III clinical trials. We mechanistically validated the top-ranked candidates in neuronal cells, identifying drug combinations with efficacy in reducing oxidative stress and safety in maintaining neuronal viability and morphology. Our neuronal response experiments confirmed several biologically efficacious drug combinations. This pipeline showed that harmonizing heterogeneous and complementary data/knowledge, including human interactome, transcriptome patterns, experimental efficacy, and real-world patient data shed light on the drug development of complex diseases.

**One-Sentence Summary:** A novel multitask deep learning method that synthesize heterogeneous biological knowledge to identify repurposable drugs for Alzheimer’s Disease.

## INTRODUCTION

Developing drugs for treating Alzheimer’s disease (AD) has been extremely challenging and costly. The total cost of developing new AD drugs, including failures, is estimated at $5.7 billion - seven times more than the cost of developing cancer medicines (Cummings et al., 2018). The most recent FDA approval of Aducanumab is an amyloid beta-directed monoclonal antibody, which aims to treat patients by reducing the buildup of β-amyloid (Office of the Commissioner, 2021). Despite showing some promising efficacy in removing β-amyloid, only patients with mild stage and amyloid burden can be benefited from the expensive treatment (estimated to cost $56,000 per year) (Fleming et al., 2020). To develop potential treatments in a timely and cost-effective manner, drug repurposing has shown good potential due to the reduced risk of drug toxicity of previously approved drugs. What is even more promising is to study combinatorial drug repositioning, which has demonstrated their potential for synergistically treating complicated diseases, including cancer (Kim et al., 2021), diabetes (Cahn and Cefalu, 2016), metabolic syndrome (Bakris et al., 2006), and cardiovascular disease (Working Group on the Summit on Combination Therapy for CVD, 2013), thanks to their ability to target multiple pathologies.

Most current AD drug repurposing studies investigate various perspectives, including drug perturbation transcriptome profiles, network pharmacology, and treatment effects in real-world patient data. Transcriptomic-based strategy compares drug-induced gene expression with gene expression in AD specimen (Siavelis et al., 2016; Sirota et al., 2011; Williams et al., 2019), which captures integrated molecular changes in different AD pathology. Network pharmacology is another approach that represents a drug’s multi-target capacity in a human interaction network (Chen and Xu, 2019; Huang et al., 2019; Pham et al., 2020). This method aims to identify hidden interactions among drugs and proteins (or disease targets) by estimating the proximity between entities in the network. Alternatively, the real-world data approach leverages large sets of patients’ drug administration data to obtain off-label indication via treatment effect estimation (Geifman et al., 2017; Zissimopoulos et al., 2017). Each strategy captures different aspects in multimodality and multiscale of the AD treatment landscape. These different approaches provide complementary evidence into potential drugs (Cheng et al., 2018). Success in a single aspect, however, cannot guarantee clinical effectiveness due to AD’s heterogeneous pathogenesis. Consequently, there is a critical need to integrate diverse evidence of transcriptomic observations, heterogeneous biological interactions, preclinical experimental results, and real-world observation to target multiple perspectives in AD drug development.

In this study, we proposed an integrated framework for AD drug repurposing. We postulated that by harmonizing drugs’ molecular profiles and chemical structures into a knowledge graph, our framework can be used for efficient and systematic identification of potentially repurposable drugs. Here, we implement a graph representation learning model that can simultaneously encapsulate the heterogeneous interactions in the graph and discriminate the molecular determinants of AD into embeddings. Subsequently, our multi-task ranking model integratively prioritizes candidates based on multiple levels of drug evidence, including drug perturbation transcriptome profiles, clinical trial history, drug efficacy in preclinical experiments, and population-based treatment effect estimation. Mechanistic validations showed our candidates’ strong efficacy in reducing oxidative stress in mouse brain cell lines. In summary, our key contribution includes:

- Developing advanced deep representation learning to integrate isolated biological knowledge and data, followed by mechanistic validation using mouse brain cell, and
- Covering comprehensive knowledge and data from molecular interactions, transcriptome, preclinical/clinical trials, and population-based treatment effect in real-world patient data.

## RESULTS

### AD knowledge graph representation

In this study, we utilized an advanced Graph Neural Network (GNN) approach to identify repurposable drug candidates in the AD knowledge graph. We first integrated biological interactions from multiple human-curated databases to build a comprehensive AD knowledge graph with 30,279 nodes and 398,644 edges. The AD knowledge graph has four types of nodes, including drugs, genes, pathways, and gene ontology, which were connected by seven different interaction types (Figure 1a, Method 1). We designed a graph autoencoder model to transform the complex relationships within the AD knowledge graph into embedding (Method 2). Underlying AD etiology is an open research area and current knowledge of AD-related interactions is likely incomplete. Previous study has shown that transfer learning from large pre-trained knowledge graph models can enhance model performance in predicting relatively limited domain knowledge such as a COVID-19 biological network (Hsieh et al., 2021). To address the incomplete knowledge of AD, we utilized transfer learning, a machine learning approach that leverages a pre-trained model to warm-start training process and improve performance, from the universal biological database representation in the DRKG network (Zeng et al., 2020) (Method 3). Additionally, we encouraged the gene embedding in the knowledge graph to distinguish high-risk AD-related genes as an auxiliary task (Method 4). To achieve these goals, we jointly optimized our model to learn the high-risk AD-related genes (node classification) and known relationships among AD knowledge representation (edge classification).

**Figure 1.**
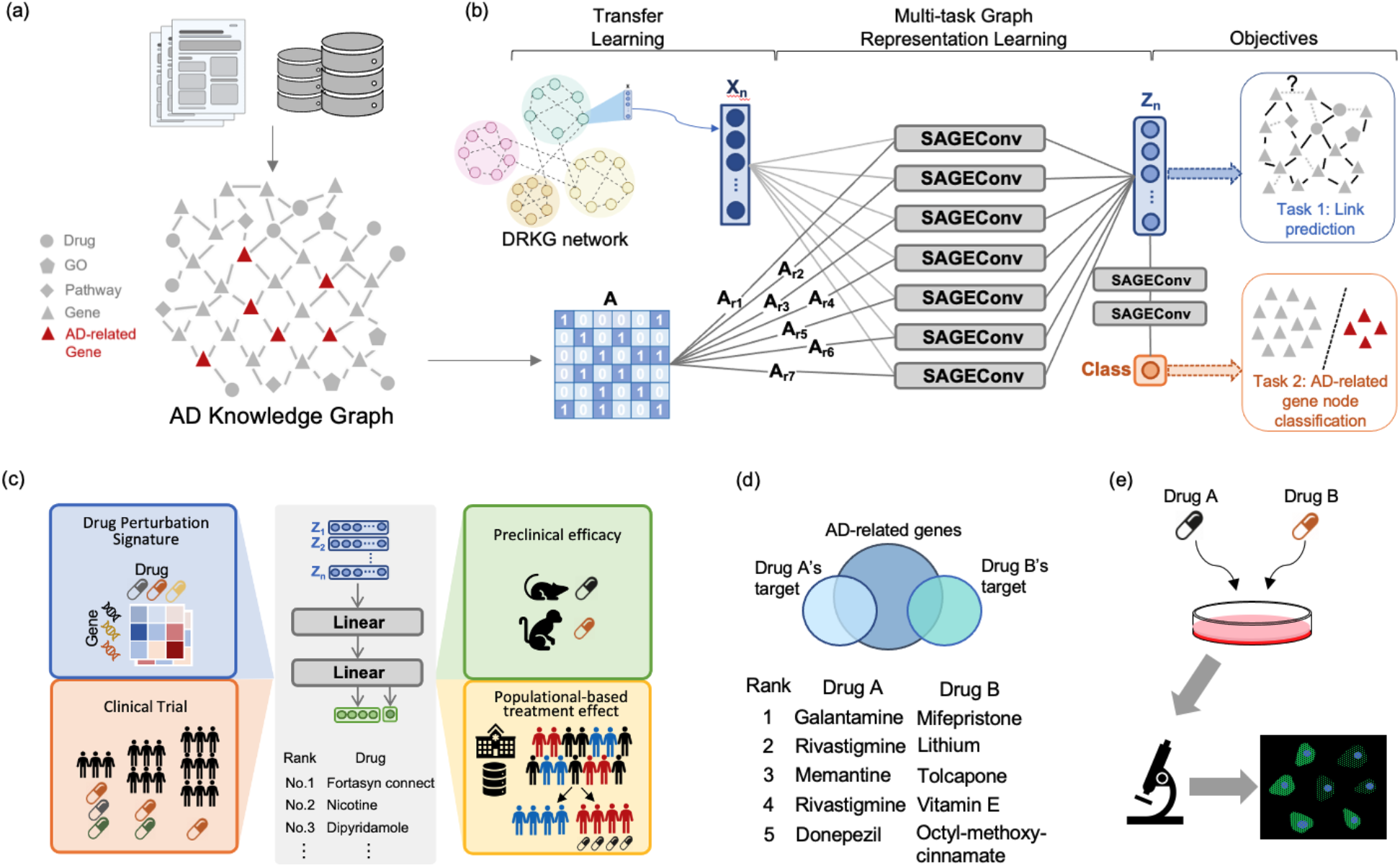
Study workflow. (a) Build the AD knowledge graph with nodes (drugs, genes, pathways, and gene ontology or GO) and edges (drug-target interaction, drug-drug structural similarity, gene-gene interaction, gene-pathway association, gene-GO association, and drug-GO association). (b) Derive the node embeddings using our multi-relational variational graph autoencoder (Kipf and Welling, 2016; Schlichtkrull et al., 2017). (c) Multi-task ranking model was used to rank drug candidates based on multi-level evidence in drug perturbation signatures, preclinical efficacies, clinical trials, and population-based treatment effects. (d) Search drug combinations satisfying complementary exposure patterns (Zhou et al., 2020) using the high-ranked drug candidates. (e) Validate drug combinations using oxidative stress assays on HT22 Mouse Hippocampal Neuronal Cell Line.

### Evaluate knowledge graph representation

After training the model to learn heterogeneous relationships in the AD knowledge graph, we evaluated how well the embeddings faithfully condense the AD knowledge. Specifically, we investigated whether the embedding could restore the known interactions in the knowledge graph with or without transfer learning (Method 5, Table 1), and visually examined the global position of the node embedding via UMAP plot (Figure 2) (McInnes et al., 2018). As a result, our knowledge graph representation achieved the area under the precision-recall curve (AUPRC) of 0.958 and mean average precision (MAP) of 0.1207 without transfer learning and the AUPRC of 0.988 and the MAP of 0.3195 with transfer learning, which are higher than universal embedding (Method 3) that achieved the AUPRC of 0.648 and MAP of 0.1013 (Table 1a). To detect any under-representation in the embeddings, we separately evaluated accuracy of each edge type. Our model consistently predicted most edge types. The Drug-GO interactions are relatively harder to predict, whereas gene-gene interactions are easier to predict (Table 1b). By ablating one edge type and measuring the edge prediction accuracy, we found that the drug-target or drug-pathway interactions are the most unique interactions that are hard to be inferred if they are excluded (Table S1). For the AD-related gene prediction task, the gene embedding was well trained to distinguish the 743 AD-related genes from other remaining genes with the area under the receiver operating curve (AUROC) of 0.947 and AUPRC of 0.583 (Table S2). Analysis showed that the model incorrectly predicted TRIOBP (probability = 0.0476) as a non-AD gene, likely due to the connection to fewer neighboring nodes and thus limited the model propagation, compared with ADAM10 (probability = 0.826) (Figure S1).

**Table 1.** Edge prediction accuracy using AD knowledge graph’s node embedding. (a) Edge prediction performance of each set of embeddings. (b) To examine how well each edge type is preserved in the node embedding, we measured the edge prediction accuracy for each edge type separately. The accuracy values for all edge types were consistent without any edge types being under-fitted. For AD knowledge graph representation without transfer learning, we initialized the node embedding with random initialization.

|  | Pretrained universal embedding | AD knowledge graph representation without transfer learning | AD knowledge graph representation with transfer learning |
| --- | --- | --- | --- |
| AUROC | 0.565 | 0.959 | 0.991 |
| AUPRC | 0.648 | 0.958 | 0.988 |
| Mean reciprocal rank (MRR) | 0.0057 | 0.0264 | 0.1112 |
| Mean Average precision (MAP) | 0.1013 | 0.1207 | 0.3195 |
| Precision@10 | 0.1235 | 0.0613 | 0.2517 |

(b)
| Which edge type to predict | Pretrained universal embedding |  | AD knowledge graph representation without transfer learning |  | AD knowledge graph representation with transfer learning |  |
| --- | --- | --- | --- | --- | --- | --- |
|  | AUROC | AUPRC | AUROC | AUPRC | AUROC | AUPRC |
| Drug - gene | 0.528 | 0.489 | 0.863 | 0.795 | 0.960 | 0.941 |
| Gene - gene | 0.987 | 0.985 | 0.936 | 0.887 | 0.991 | 0.984 |
| Drug - GO | 0.604 | 0.554 | 0.657 | 0.578 | 0.889 | 0.895 |
| Gene - GO | 0.613 | 0.593 | 0.925 | 0.912 | 0.985 | 0.974 |
| Drug - pathway | 0.282 | 0.376 | 0.989 | 0.988 | 0.998 | 0.995 |
| Drug - drug<br>structural<br>similarity | 0.897 | 0.867 | 0.981 | 0.985 | 0.994 | 0.990 |
| Gene -<br>pathway | 0.397 | 0.459 | 0.590 | 0.639 | 0.956 | 0.947 |

**Figure 2.**
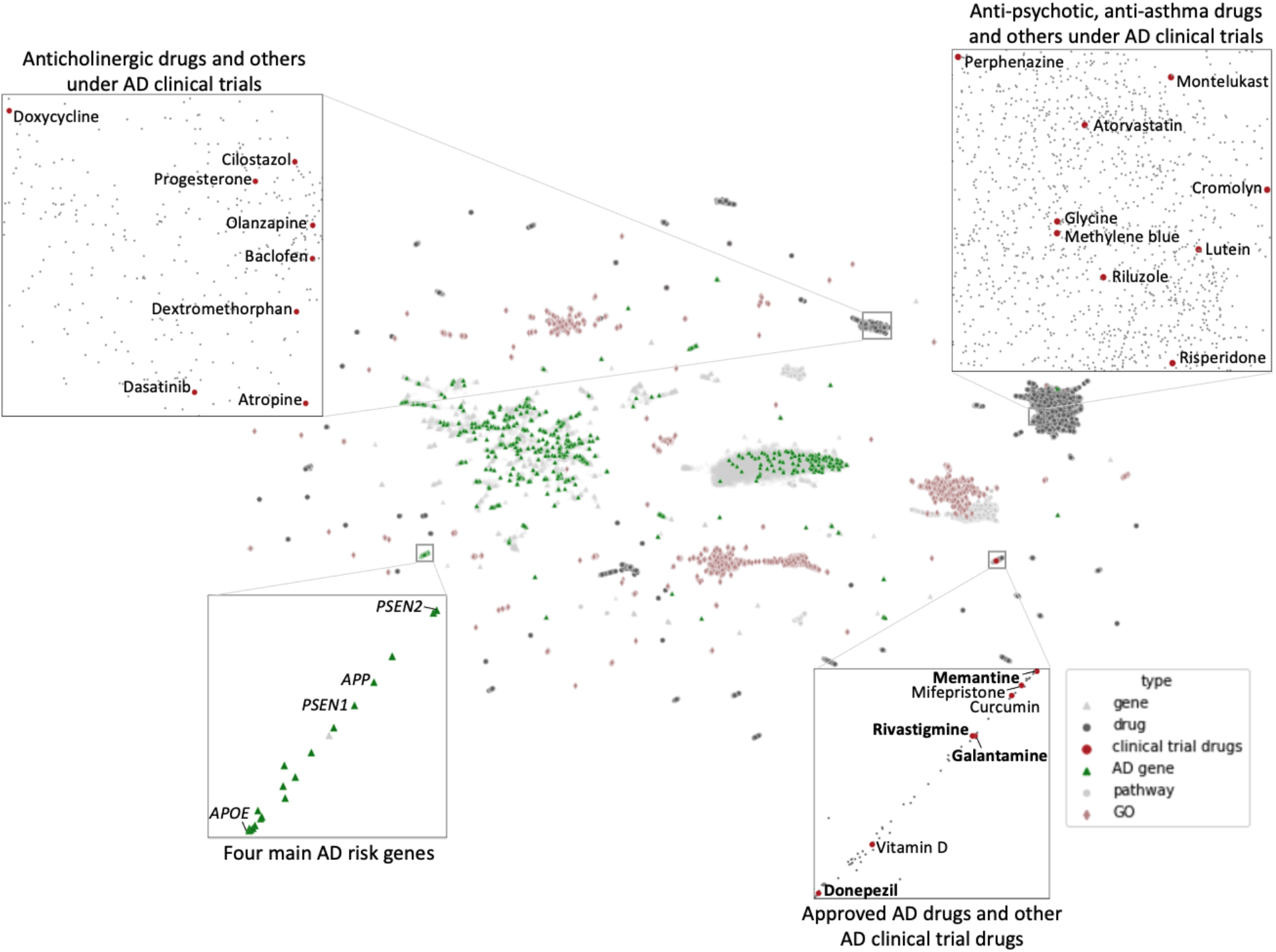
UMAP plot for lower-dimensional projection of node embedding. Nodes with similar embeddings are adjacent in the UMAP plot. Four current FDA-approved AD drugs were indicated in bold text. Four main AD risk genes (green triangle) and undergoing clinical trial drug (red circle) clusters were highlighted in subplots. Genes, which are in gray triangles, were mixed with drugs, which are in the black rounds. Gene ontologies (pink diamond) and pathways (light grey circle) are adjacent to genes and drugs.

Visualizing the embedding via UMAP, we found that the overall node distribution is generally dispersed throughout the plot rather than clustered with the same node type (Figure 2). Undergoing clinical trial drugs are aggregated in three main groups, each with similar therapeutic or pharmacological categories, implying the node embedding well reflects the contextual information (Figure 2). In all, the high accuracy in masked interaction prediction and AD-related gene prediction, together with global distribution in UMAP plot, implies that the node embedding faithfully captures the local and global network topology of the AD knowledge graph regardless of edge types, with the help of universal embedding that provides complementary information to restore the masked AD knowledge. This implied the potential applicability of our derived embeddings for drug repurposing tasks.

### Identifying drug candidates using multi-level evidence

Most prior studies have identified repurposable drugs based on missing edge prediction between drug and target in GNN (Mohamed et al., 2020; Zitnik et al., 2018). However, this approach is heavily dependent on the reliability of AD-related high-risk genes, which is still an open research question. Instead, our approach is a “reverse engineering” approach that utilizes drugs with nontrivial evidence in preclinical/clinical stages and identifies similar drugs in terms of AD biology. After synthesizing the heterogeneous AD knowledge into the embedding, we prioritize repurposable drugs by taking four categories of evidence as labels, including transcriptomic reversed patterns, mechanistic efficacies, population-based treatment effects (Table S3), and clinical trials (Method 6). Our multi-task ranking model utilizes a pairwise ranking loss to prioritize repurposable drugs (Method 7). As a result, the ranking model accuracy was AUROC=0.910 and AUPRC=0.472 to predict drugs with at least one of the drug efficacies labels. We presented the top-ranked drugs in Table S4 (precision at top 600=0.42) and highlighted the top 10 in Table 2.

**Table 2.**
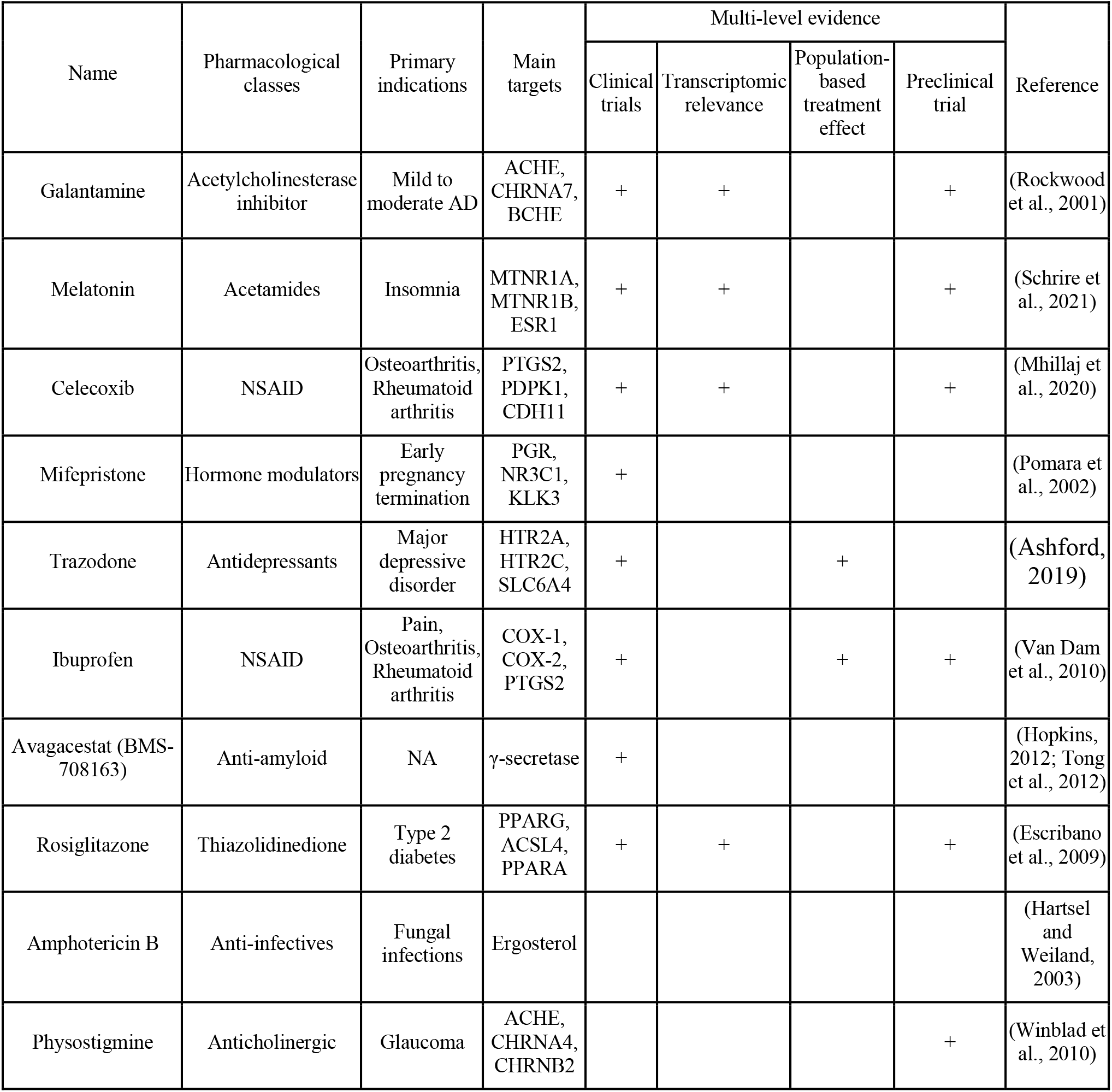
Selected promising drugs with supporting evidence and literature. +: positive evidence, blank: not investigated.

### Identifying drug combination using complementary exposure pattern analysis

As indicated by the complexity of the AD knowledge graph, using single drugs to treat AD might result in limited effects. To improve treatment efficacy, we identified potential drug combinations from the top ranked drugs (Table S4) and existing FDA-approved AD drugs. Four conventional drugs treat symptoms in AD – rivastigmine, galantamine, donepezil, and memantine, are generally effective in two main mechanisms of action, cholinesterase (AChE) inhibitors or *N*-methyl D-aspartate (NMDA) antagonists. These drugs are intended to maintain or stabilize neuronal function, management of behavioral symptoms, and slow down the rate of memory loss by regulating neurotransmitters. In general, AChE inhibitors are recommended for the treatment of mild-to-moderate AD, whereas NMDA antagonist is prescribed to treat moderate-to-severe AD (Plascencia-Villa and Perry, 2021). Unfortunately, after high-dose exposures to these drugs, patients often experience various CNS and metabolic side effects, including nausea, loss of appetite, weight loss, headache, and confusion, etc. To maximize effect while attempting to minimize adverse reactions by reducing dosage of the conventional medications, we proposed to discover alternative treatments for AD by combining the FDA-approved AD drugs with the top-ranked drugs identified in our computational model.

To identify drug combinations with synergistic interactions without degradation in safety, we leveraged the “Complementary Exposure Pattern” (Method 8) analysis, which has been successfully applied to drug repurposing in hypertension (Cheng et al., 2019) and COVID-19 (Hsieh et al., 2021). The theory indicates that a drug combination is therapeutically effective if the targets of the drugs hit the disease module without overlap (Cheng et al., 2019). Table 3 summarized the identified drug combinations in our top 30 drugs, and the amount of their common targets. A total of 10 combinations satisfied the criteria, where the combination of Galantamine and Mifepristone has the widest coverage to the AD module. The efficacies of these combinations were further validated using neural cell models.

**Table 3.** Drug combinations that satisfy the complementary exposure pattern from the top 30 drugs.

| Drug A | Drug B | # AD target A hits | # AD target B hits | # AD target A or B hit |
| --- | --- | --- | --- | --- |
| Galantamine | Mifepristone | 7 | 90 | 97 |
| Rivastigmine | Lithium | 7 | 59 | 66 |
| Memantine | Tolcapone | 10 | 28 | 38 |
| Rivastigmine | Vitamin E | 7 | 29 | 36 |
| Donepezil | Octyl-methoxycinnamate | 25 | 8 | 33 |
| Galantamine | Diclofenac | 7 | 26 | 33 |
| Rivastigmine | Diclofenac | 7 | 26 | 33 |
| Galantamine | Caffeine | 7 | 25 | 32 |
| Donepezil | Manidipine | 25 | 6 | 31 |
| Memantine | Octyl-methoxycinnamate | 10 | 8 | 18 |

### In vitro validation on safety, tolerability, and neuronal responses of drug combinations

To examine the potential efficacies of our predicted drug combinations, we mechanistically validated the top-ranked drug combinations by measuring cytotoxicity and oxidative stress on hippocampal neurons (Method 9, Figure 1f). Our results suggest significant efficacy of reducing reactive oxygen species (ROS) levels by multiple drug combinations, implying the potential applications of combining existing and novel drug candidates for AD.

Memantine functions as an antagonist of glutamatergic NMDA receptors, type 3 serotonin (5HT3) receptor, and nicotinic acetylcholine receptor (nAChRs), allowing neuronal availability of neurotransmitters and improving cognition and behavior in moderate-to-severe AD patients. In combination with Tolcapone, an FDA-approved catechol-O-methyltransferase inhibitor used in the management of Parkinson’s disease to increase levels of peripheral dopamine, we observed safety and tolerability in cholinergic neurons during the dose- and time-course treatments. The combination can reduce the production of ROS by 5.6±2.2%. In the presence of Aβ, Mem+Tolca was able to reestablish oxidative stress to basal levels, whereas the single Mem treatment presented a 5.5±2.7% increase (Figure 3). Memantine is usually administered in combination with Donepezil, but in combination with Tolcapone, the drugs could have significant benefits on global and functional outcomes in AD subjects (Apud et al., 2007; Fremont et al., 2020).

**Figure 3.**
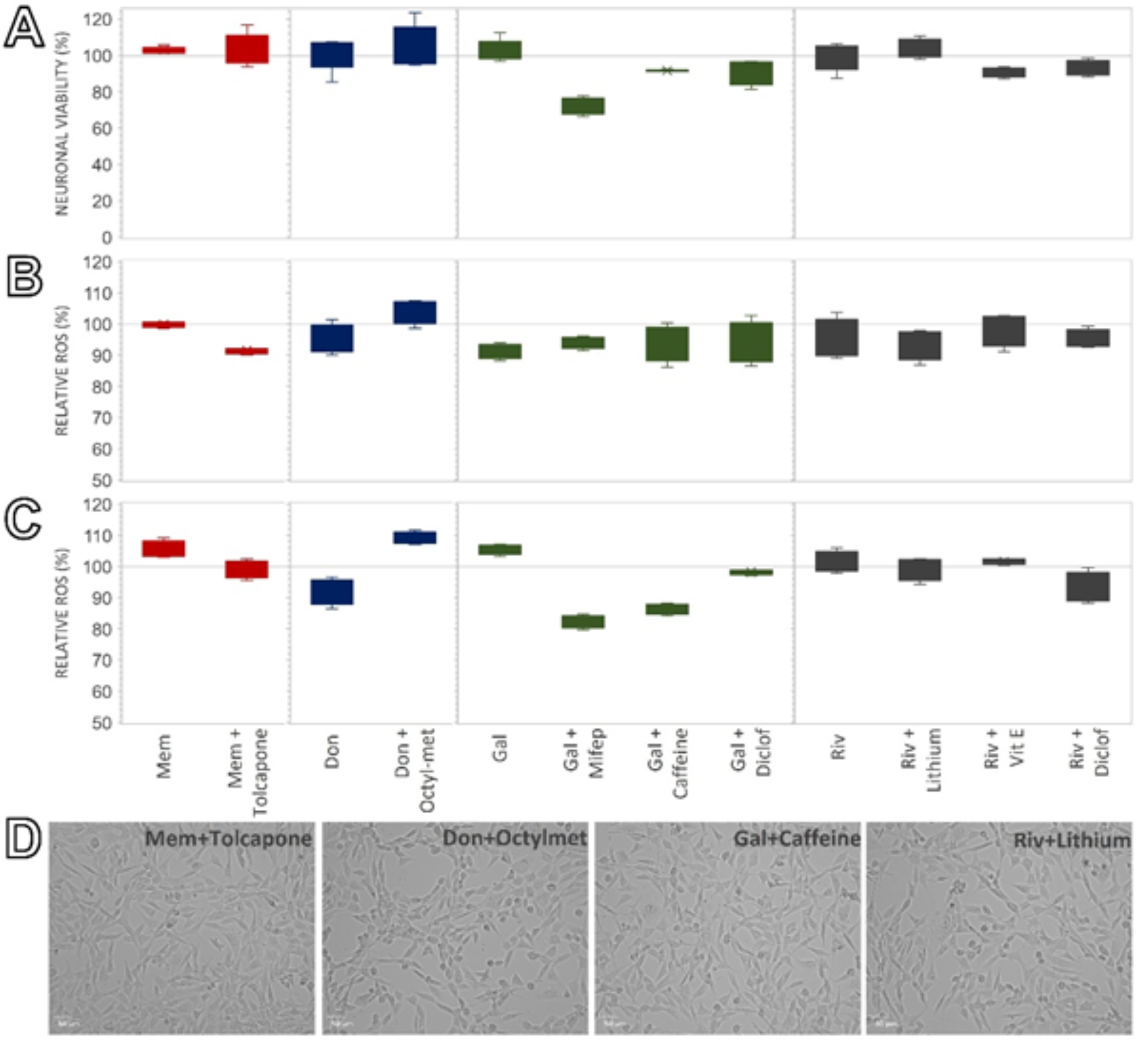
Neuronal responses to drug combinations. (A) Neuronal viability after 24 h with 50 μM of drug combinations. (B) Quantification of intracellular oxidative stress (DCFDA fluorescence) after 24 h with 50μM of drug A + 50μM of drug B. (C) Quantification of intracellular oxidative stress (DCFDA fluorescence) of neurons dosed with 10 μM Aβ, after 24 h with 50μM of drug A + 50μM of drug B. All data sets were normalized to neurons dosed with vehicle (DMSO) and the control was represented as a horizontal line at 100% across all the treatments. (D) Live-cell brightfield imaging of neurons dosed for 24 h with 50μM of drug A + 50μM of drug B.

Donepezil, an AChE inhibitor that allows an increase in acetylcholine in cholinergic neurons, has been used for 25 years to delay cognitive decline and symptoms of AD. Among our top-ranked drugs, the combination of Don+Octyl-methoxycinnamate showed high tolerability and safety. Functionally, Donepezil effectively reduced the amount of ROS by 6.82-7.98%, but its combination with Octyl-met showed a slight increased by 4.3-9.1% (Figure 3), this could be related to the limited solubility of this active compound and that it is usually employed in topical formations.

Galantamine is a weak AChE inhibitor and allosteric potentiator of acetylcholine receptors, intended to treat the progression of symptoms, memory loss, and thinking ability in mild-to-moderate AD. Gal prevents acetylcholine processing and stimulation of its release by nicotinic receptors. Our results showed that combining Galantamine with Mifepristone, Caffeine, or Diclofenac, can lead to a reduction in oxidative stress responses by 5.8-6.3%. These neuroprotective effects were more evident in the neurons dosed with Aβ, decreasing ROS production at 17.7% in Gal+Mifep, 13.5% in Gal+Caffeine, and 2% in Gal+Diclofenac, whereas single Galantamine treatment presented a 5.5% increase in neuronal oxidative stress (Figure 3). Previous studies found that the neuroprotective effect of Mifepristone is related to preventing stress-induced responses, protecting neurons from undergoing programmed cell death (apoptosis), and increasing AMPA receptor expression (Ghoumari et al., 2006), while Mifepristone may also promote rapid repair of the synaptic alterations in the hippocampus (Wu et al., 2007). Alternatively, Caffeine can activate noradrenaline neurons and the release of dopamine, counteracting the development of cognitive impairment as an antioxidant (Londzin et al., 2021). On the other hand, Diclofenac, an anti-inflammatory drug targeting voltage-dependent K^+^ channels, also has neuroprotective effects in neurons and showed favorable outcomes of reducing the AD risk in veterans (Naeem et al., 2019; Stuve et al., 2020).

Acting as an AChE and butyrylcholinesterase inhibitor, Rivastigmine is used to treat memory loss and improve thinking abilities in AD patients. The tolerability and safety of Rivastigmine alone or in combination with Lithium, Vitamin E, and Diclofenac showed well tolerance in cholinergic neurons (Figure 3). Neuronal responses indicated that these active compounds can effectively reduce oxidative stress in neurons by 4.8-6.2%. In the presence of Aβ, Rivastigmine, Riv+Lithium, and Riv+Vit E showed similar levels of ROS reduction, whereas Riv+Diclof can further reduce 6.8% of intracellular ROS (Figure 3). Lithium, currently under Phase III clinical trials for treating AD, can reduce excitatory neurotransmitters and increase GABA, and prevent cognitive and functional impairment in mild dementia (Forlenza et al., 2019; Malhi et al., 2013). Vitamin E (tocopherol), a neuroprotectant with antioxidant properties, is under clinical trials (Phase II/III) to determine its effects on clinical progression of AD (Sen et al., 2004). Taken together, this study successfully identified several drug candidates with high potential of treating AD and have optimistic therapeutic values, and numerous drug combinations suggested by our pipeline also showed promising efficacies in reducing oxidative stress in neural cell models.

## DISCUSSION

The objective of this study was to provide proof-of-concept hypotheses on drug combinations to treat AD, utilizing knowledge graph embedding of heterogeneous interactome and multi-task learning of complementary evidence on drug efficacies. Our AD knowledge graph embedding boosted by pretrained universal pharmacological embedding integrates not only the human interactomes but also millions of interactions in public biomedical literature. Our multi-task learning of AD-related genes and multi-level drug efficacy aims to close the gap between the scattered knowledge in AD research.

Our methodology tackles important challenges to harmonize fragmented multimodal data. Recently, several prior works have used the graph neural network approach for drug development, either for general purpose (Ioannidis et al., 2020.; Mohamed et al., 2020), for rare diseases (Sosa et al., 2020), or for specific diseases including COVID-19 (Gysi et al., 2021; Hsieh et al., 2021; Zeng et al., 2020) and AD (Sügis et al., 2019). Our methodological strength over the prior approach include (1) transfer learning of universal embedding to integrate broader modality to address the incomplete knowledge, (2) multi-task learning for multimodal interactions, AD-related genes, and multi-level drug efficacy to mitigate the lack of established therapeutic targets, and (3) *in vitro* validation of the computational model via hippocampal neuron cells.

Drug combinations that we validated are less toxic (FDA-approved drugs) and economically viable (generic drugs). AD is multifactorial and the use of multi-target therapeutic interventions addressing several molecular targets seems to be an alternative approach to modify the course of AD progression (Kabir et al., 2020). Different clinical studies demonstrate that a combination therapy is more efficient than monotherapy, especially in mild-to-moderate AD to slow down the rate of cognitive impairment. Our integrative approach has identified promising drug combinations adjuvant to existing FDA-approved AD drugs. All drug combinations we identified are available in generic formulations, making them economically viable and available as an alternative to patented formulations with a high monthly cost (>500 USD).

Our study has several limitations. Instead of a mechanism-based approach that identifies therapeutic targets first and identifies drugs interacting with them, our approach relies on historical or data-driven evidence, such as estimated drug efficacy (transcriptomic reversed patterns, population-based treatment effect) and established efficacy (preclinical or clinical trials). If this data-driven drug efficacy is biased, our model’s drug ranking can be also biased. For example, drugs targeting amyloid beta are much more studied than drugs targeting other pathologies, thus our model has a potential risk to trap in the circularity of failed drugs targeting amyloid beta. Population-based treatment effects in real-world data can partly mitigate the bias in historical research data toward amyloid-beta. Finally, in vitro patterns do not guarantee in vivo efficacy or treatment outcomes in patients due to various reasons, including diverse genomics profiles in AD transgenic mouse models, variation of PK/PD and bioavailability in the human body, etc. However, our contribution is to efficiently and comprehensively refine a list of candidates from a huge set of compounds, accelerating the costly early-stage drug screening step in the drug development process. Our computational predictions and in vitro validations provided a list of highly promising drug combinations and opened future research opportunities.

Despite the limitations, our drug repurposing strategy has a broad impact not only on AD but also on complex diseases, in which complex mechanisms of actions and therapeutic targets are an open question. The well-known deficiency of knowledge and insufficiency of data in AD research, when studied separately, prevent us from capturing the complex AD’s heterogeneous pathogenesis. Our pipeline integrates complementary information at multiple stages (such as transcriptomic patterns, preclinical efficacy, clinical partial effectiveness, and population-based treatment effect) to better identify promising drug combinations. Hypotheses generated from this integrative approach may shed light on complex disease drug development.

## STAR METHODS

### RESOURCE AVAILABILITY

#### Lead contact

Further information and requests for resources and reagents should be directed to and will be fulfilled by the lead contact, Yejin Kim.

### MATERIALS AVAILABILITY

This study did not generate any new reagents.

### DATA AND CODE AVAILABILITY

- All interaction data used in this study are publicly available from CTD, STRING, Agora, ClinicalTrial.gov, and AlzPED.
- Experimental results are available in supplementary materials.
- All original code has been deposited at https://github.com/freshnemo/AD-KG, and is publicly available as of the date of publication.

### MATERIALS AND METHODS

#### 1. Build AD knowledge graph

To construct a high-quality AD knowledge graph that represents the disease-related biological interactome, we extracted and harmonized three major data sources that contain the following interaction types: (1) Universal protein-protein interactions from STRING (Szklarczyk et al., 2017); (2) Interactions between genes, drugs, GO, and pathways from the Comparative Toxicogenomics Database (CTD) (Davis et al., 2017, 2009); (3) Drug-drug associations based on four types of structural similarity fingerprints. Genes were annotated to their Entrez IDs assigned by the National Center for Biotechnology Information (NCBI) database. Details of each relation type will be described below.

##### Drugs and genes

We collected precompiled drug-gene interactions from CTD, including 6,543 unique FDA-approved drugs or experimental compounds, as well as 16,997 genes that interact with those drugs. 119 drugs have direct evidence reporting either the drugs’ relevance to AD etiology (e.g., Streptozocin) or treatment (e.g., Donepezil). The remaining drugs are from literature reporting the potential efficacy of the drugs. 117 genes have direct evidence reporting either the gene’s relevance to AD etiology (e.g., APOE) or treatment (e.g., LEP). The remaining genes are from literature reporting the potential relevance of the genes to AD. The 110,637 drug and target gene interactions are from literature with cell-based or animal-based evidence.

##### Gene and gene

We retrieved gene-gene interaction from the STRING database, which collected seven types of evidence for protein-protein interaction: biological experiment, pathway inference, text mining, gene co-expression, neighborhood genes in chromosomes, fusion, and co-occurrence (Szklarczyk et al., 2017). We selected 123,738 protein-protein interactions that have >95% combined confidence scores across the seven points of evidence.

##### AD-related genes

To highlight important genes related to disease progression or treatment strategies in the AD knowledge graph, we extracted high-confident AD-related genes from (1) Agora’s nominated gene list, identified and validated by researchers from the National Institute on Aging’s Accelerating Medicines Partnership in Alzheimer’s Disease (AMP-AD) consortium (https://agora.adknowledgeportal.org/); (2) CTD’s gene list, with direct evidence of being a biomarker or a therapeutic target. In total, we obtained 743 AD-related genes, which were utilized in the node classification task (see **Method 4**).

##### Drug and drug structural similarity

A drug with a similar structure usually accompanies similar targets. To best represent each compound’s chemical structure, we utilized four types of fingerprints: atom-pair fingerprints (Carhart et al., 1985), MACCS fingerprint (Durant et al., 2002), Morgan/Circular (Morgan, 1965), and Topological-torsion fingerprints (Nilakantan et al., 1987). Sørensen–Dice similarity coefficients were computed between every two drugs (or compounds) using RDkit (version 2021.03.4) (Landrum, 2010). For each type of fingerprint, we calculated the *z*-score of the Sørensen–Dice coefficient to obtain pairwise compound similarity and defined compound pairs with *z*-score>=3 to be structurally similar. In total, we determined 37,669 compound pairs that are structurally similar to each other.

##### Pathway and AD-related genes

For each AD-related gene, CTD linked them with their involved pathways from Reactome (Fabregat et al., 2018) and KEGG (Kanehisa et al., 2017) databases. Various pathways of a multifactorial disease like AD can display the diverse etiology and mechanisms of the disease. Querying “Alzheimer’s Disease” from CTD, we extracted 1,778 gene-pathway interactions containing 678 unique pathways.

##### Pathway and drugs

Among 6,543 drugs of interest, their drug targets from CTD were used to query related Reactome (Fabregat et al., 2018) and KEGG (Kanehisa et al., 2017) pathways and computed enriched drug-pathway associations using Enrichr (Subramanian et al., 2005). For each drug, enriched pathways for all its target genes were identified using a threshold of p-value < 0.05. Drugs and their enriched pathways were connected as drug-pathway interactions. There are 110,637 drug-pathway interactions, including 1,622 pathways in our knowledge graph.

##### GO and AD-related genes

Gene ontologies are structured and controlled vocabularies that represent each gene’s participating biological processes, molecular functions, and cellular components. We extracted gene ontologies for AD-related genes from CTD to construct gene-GO edges. A total of 7,368 gene-GO interactions were collected between 89 AD risk genes and 2,965 unique GO terms.

##### GO and drugs

CTD also inferred associations between GO and certain drugs under Alzheimer’s Disease. Drug-GO interactions were established based on either a combination of curated drug-GO interactions and drug-AD interactions, or a gene-GO annotation in AD or both. These expanded our AD knowledge graph with 6,817 drug-GO interactions, including 2,377 unique GO terms and 108 unique drugs.

#### 2. Knowledge graph representation

Graph neural network (GNN) is one field of deep neural networks that derive a vectorized representation of nodes, edges, or whole graphs. Adopting GNN into the biomedical network facilitates the integration of multimodal and complex relationships. The graph node embedding can preserve the node’s local role and global position in the graph via iterative and nonlinear message passing and aggregation. It learns the structural properties of the neighborhood and the graph’s overall topological structure (Hamilton et al., 2017), thus allowing us not only to restore the known interactions but also infer unknown interactions. The knowledge graph representation that contains such putative interactions can help us find new indications of drugs (Gaudelet et al., 2021). Recently GNN has demonstrated a great advance in predicting hidden interactions (e.g., PPIs, drug-drug adverse interactions, and drug-target interactions) and the discovery of new molecules (Mohamed et al., 2020; Zitnik et al., 2018). GNN is also used to derive representation from a graph with multiple types of relations and nodes (i.e., heterogeneous network or knowledge graph). The Drug Repurposing Knowledge Graph (DRKG) (Zeng et al., 2020) uses the knowledge graph representation to capture whole topological relations from seven biomedical databases (Percha and Altman, 2018; Zeng et al., 2020). The knowledge graph includes 97,238 nodes (from 13 node types including gene, molecular function, pathway, disease, symptom, anatomy, cellular component, compounds, side effect, ATC, and pharmacologic class) and 5,874,261 interactions (from 107 edge types) from DrugBank (Law et al., 2014), HetioNet (Himmelstein et al., 2017), GNBR (Percha and Altman, 2018), STRING (Szklarczyk et al., 2018) and IntAct (https://www.ebi.ac.uk/intact/). Their representation offers a general and universal embedding of these entities, which can further enrich other domain-specific knowledge representations.

In this study, we developed the knowledge graph representation model by customizing the deep variational graph neural autoencoders (VGAE) to incorporate multiple types of relations or edges (Figure 1b) (Kipf and Welling, 2016). The self-supervised graph autoencoder method encodes the nodes into a latent vector (embedding) and reconstructs the given graph structure (i.e., graph adjacency matrix) with the encoded latent vector. The variational graph approach can encapsulate the complex network into a probabilistic distribution (rather than deterministic vector representation) for nodes considering the uncertainties of our knowledge for AD; therefore, it alleviates the overfitting/underfitting issues due to partial knowledge. We customized the VGAE to incorporate the different types of edge by setting different weight matrices for each type of edge. This multi-relational approach respects the different propagation for each interaction type. Since our objective is to derive node embedding that reflects the overall graph topology, the multi-relational VGAE model was trained to reconstruct the missing interaction using the node embeddings as an autoencoding manner. For the optimization, in the message-passing step, each node (entity)’s embedding is iteratively updated by aggregating the neighbors’ embedding, in which the aggregation function is a mean of the neighbor’s features, concatenation with current embedding, and a single layer of a neural network on the concatenated one. PyTorch Geometric was used for model implementation (Fey and Lenssen, 2019). The model structure was (1×400) → Graph convolution to (1×256) → RELU → Dropout → Summation of multiple edge types → Batch norm → Graph convolution to 1×128 (mean) and 1×128 (variance). We randomly split the knowledge graph edge (i.e., positive edges) into 90% for training and 10 % for testing, and generated the same number of negative edges from which the positive edges will be predicted.

#### 3. Transfer learning from a comprehensive drug knowledge graph

We boosted the node embedding’s biological relevance in the AD knowledge graph with millions of interactions from public biomedical literature by transfer learning. Transfer learning is to transfer knowledge from previously learned universal models to domain-specific models. By transferring and fine-tuning the universal knowledge, the AD network embedding can respect universal pharmacological interactions while prioritizing AD-related interactions. Injecting this universal knowledge into the AD domain is critically important in addressing the uncertainty in AD knowledge. To achieve this task, we extracted the DRKG network (Zeng et al., 2020), which contains the universal embedding of 15 million pharmacological entities across 39 different types of interactions among genes and compounds from large biomedical databases including DrugBank, HetioNet, GNBR, STRING, IntAct, and DGLdb (Ioannidis et al., 2020). We avoided directly incorporating the edges of the DRKG network in our AD knowledge graph since the DRKG is comprehensive in breadth but shallow in depth. DRKG’s network contains less accurate interactions (e.g., the same drug encoded in different drug nodes due to ID mismatching) that can be diluted into our small but specific AD knowledge graph.

Pretrained embedding for drugs, genes, pathways, and functional ontology were retrieved from DRKG and used as initial node features in our model, and fine-tuned them to the AD knowledge graph. When nodes in the knowledge graph were not matched to entities in the universal embedding, we learned the embedding as parameters (e.g., one hot to embedding vectors).

#### 4. Node classification to differentiate AD-related genes in the knowledge graph

To leverage auxiliary information on known AD-related high-risk genes, we added a node classification task in our graph neural network model to differentiate the 743 AD-related genes (from Method 1) from the rest of genes in the graph. We hypothesized that one can predict AD-related genes using gene interaction with other genes, GO, and pathways, and the node classification task was to predict whether a gene is one of the AD-related genes using the gene representation in the AD knowledge graph. The node classification consists of two graph convolution layers (GraphSAGE) that sample a node’s adjacent neighborhoods and aggregate their representation. The dimensions of those two convolution layers were Graph convolution (128, 64) → RELU → Dropout → Graph convolution (64, 1). We split the gene nodes into 90% for training and 10% for testing. As the labels are not balanced (only 743 AD-related genes out of 16,997 genes), we randomly split the gene nodes into 90% for training and 10% for testing while maintaining the ratio of AD gene and non-AD gene. This AD-related gene classification task was jointly optimized with the relational VGAE so that the AD knowledge graph representation can distinguish AD-related genes from all other genes. We used PcGrad (Yu et al., 2020), a multi-task optimization technique that resolves conflict among multiple tasks’ gradients by normal vector projections, to achieve a better local optimum.

#### 5. Validating the quality of knowledge graph representation

We evaluated the quality of AD knowledge graph representation by checking whether the learned node embedding can restore the known edges. The edge prediction task is to predict whether an edge exists between head and tail nodes using the two nodes’ embeddings. The edge prediction task’s accuracy will be high if the node embedding encapsulates the edge information faithfully. For evaluation, we randomly set aside 10% of the edges for each edge type (Table S1). We sampled negative (or fake) but plausible edges at 1:5 ratio that do not exist in the knowledge graph by replacing tail nodes to other random nodes within the same node types and predicted whether a given edge is real or fake (Kipf and Welling, 2016). The model was trained using all edge types and evaluated separately for each edge type. We also ablated one edge type and observed how one edge type contributed to predicting all the edge types (Table S1). This experiment allowed us to determine which interactomes are unique and hard to be replaced or implicitly inferred by other edge types. The edge prediction accuracy was measured as general accuracy measure – the area under the receiver operating curve (AUROC) and the area under the precision-recall curve (AUPRC) and ranking accuracy measure – mean reciprocal rank (MRR), mean average precision (MAP), and precision at top *k*., considering the imbalance of positive and negative samples. For the node classification task to distinguish between AD-related genes and remaining genes, we also measured the classification accuracy using the AUROC and AUPRC.

#### 6. Identification of multi-level evidence for drug efficacy

Next, we prioritized 6,543 drug candidates that are similar to drugs with historical evidence. AD is a complex disease and its effective therapeutic targets are largely unknown, thus drugs interacting with a few targets can have limited efficacy (Oset-Gasque and Marco-Contelles, 2018). A holistic evidence may alternatively capture the efficacy in the systemic and complex disease. We believe the most desirable drug candidates would have multiple levels of complementary evidence to support their efficacy, such as post-treatment transcriptomic patterns, mechanistic efficacy in preclinical animal models, and epidemiological treatment effects in real-world patient data. We identified each evidence as follow:

##### Transcriptomic reversed patterns

We examined reversed patterns of the AD model’s genetic signature and drug-induced gene signature using gene set enrichment analysis. We leveraged a recent study (Williams et al., 2019) and identified 149 drugs with significant transcriptomic reversed patterns. Among the 149 drugs, 76 drugs were matched to our 6,543 drug candidates.

##### Mechanistic efficacy in preclinical models

Alzheimer’s Disease Preclinical Efficacy Database (AlzPED) accumulated pre-clinical evidence for effective AD drugs. We identified 338 drugs with significant efficacy from 791 animal-based studies targeting various pathologies including amyloid-beta, tau, and neuroinflammation. Among the 338 drugs, 189 drugs were matched to our 6,543 drug candidates.

##### Population-based treatment effect

We estimated drugs’ population-based treatment effect as real-world evidence of drug efficacy. Healthcare claim data is a unique resource to investigate treatment effects of prescribed drugs on reducing AD onset risk during the preclinical stage of AD. We used Optum’s de-identified Clinformatics^®^ Data Mart subscribed by UTHealth. This billing-purpose claim data is inherently noisy and sparse, thus requires careful data preparation (e.g., setting observation windows to exclude subjects who are not old enough to have AD onset, identifying AD onset via billing-purpose diagnosis and medication codes, grouping high-dimensional diagnosis codes into clinical comorbidities as confounding variables). We followed the data preparation process in our previous work (Ling et al., 2021) based on target trials (Hernán and Robins, 2016). We calculated the average treatment effect among treated (ATT) using inverse probability of treatment weighting (IPTW) (Pearl, 2014, 2000). See details at Supplementary Method 1. We identified 126 drugs with positive treatment effects (i.e., ATT<0 to reduce AD onset). Among the 126 drugs, 101 drugs were matched to our 6,543 drug candidates.

##### Clinical trials terminated at Phase II/III

Many drugs in the failed clinical trials passed several phases and showed non-trivial response rates (e.g., good response on certain subpopulations). We hypothesize individual drugs (focusing on different targets) from previously failed clinical trials respond differently on sub-populations, and therefore, cocktails of complementary drugs might cover a larger population. Also, because drug testing in clinical trials is partially efficacious in treating AD, a drug that is similar to these trial drugs (and their combinations) can have potential efficacy too. We collected 294 drugs from 2,675 interventional clinical trials for AD from ClinicalTrial.gov. Among the 294 drugs, 210 drugs were matched to our 6,543 drug candidates.

#### 7. Prioritizing repurposable drugs with multiple evidence

After collecting multi-level evidence of drug efficacies from Method 6, we built a multi-task ranking model to prioritize drugs from AD knowledge graph representation. To build a ranking model that takes into account four categories of evidence simultaneously, we adapted a neural network model with a pairwise ranking loss for each evidence (Kim et al., 2019; Rendle et al., 2012). For each task, the positive samples to prioritize were the drugs with the positive evidence, while negative samples were the remaining drugs in the AD knowledge graph. A fifth task was added to predict whether a drug has any evidence or not (has evidence = 1; no evidence = 0). Again, we utilized PcGrad (Yu et al., 2020) to handle the gradient inconsistency in the multi-task optimization. The architecture was two fully connected layers (with the size of 128→128→5) with residual connection, nonlinear activation (ReLU), dropout, batch norm in the middle, and the optimization loss (four pairwise ranking losses). For evaluation, we validated the ranking model by evaluating whether the positive drugs (drugs fall into any category of evidence) are ranked high. We measured the accuracy of the drug ranking model using the AUROC and AUPRC of top *k* drugs with 50% training and 50% test cross-validation. We purposely set the portion of the training set lower because the four categories of drug evidence are not our sole “gold standard” to prioritize drugs.

#### 8. Drug combination identification

To identify efficacious drug combinations from top-ranked drugs, we applied the Complementary Exposure Pattern (Cheng et al., 2019) analysis, which hypothesized that “a drug combination is therapeutically effective only if the targets of the drugs both hit the disease module, but they target a separate neighborhood”. We searched the drug combinations within the top-ranked drugs. We counted the number of genes in the AD module that a drug combination hits, where the drug combination’s targets are disjoint.

#### 9. In vitro validation on safety, tolerability, drug combinations

##### Neuronal culture

Hippocampal neurons (HT-22) were cultured in DMEM (Sigma-Aldrich #D6429) supplemented with 10% FBS, and 1X Antibiotic-Antimycotic (#30-004-CI, Corning). Neurons were subcultured every 3-4 days at approximately 75-80% of confluence and kept in a T-75 flask in a 5% CO2 incubator at 37°C. For bioassays, the neurons were detached with 0.25% trypsin-EDTA (Sigma-Aldrich #T3924), pelleted at 300g, and diluted in fresh DMEM. Cells were counted with LUNA-II automated cell counter (Logos) and seeded in 96-well-plates at 5000 cells/well the day before. Morphology was monitored with an inverted optical microscope (Olympus CKX31).

##### Drug combinations

All the drugs were ACS grade or higher purity, prepared fresh immediately prior to use in the bioassays. Stock solutions of the drugs were prepared by dissolving in DMSO (Sigma-Aldrich #) at 50 mM. Each active compound was then diluted with DMEM to 200, 100, 50, and 25 μM. As a control, we employed DMEM with DMSO.

##### Cytotoxicity and oxidative stress responses

Cell Counting Kit-8 (Dojindo, #CK04) was used to determine cell viability, proliferation, and cytotoxicity, measuring the absorbance of water-soluble tetrazolium salt (reduced WST-8) at 450 nm. Neurons were subcultured in 96-well plates and dosed with the active drugs diluted in DMEM, incubated at 37°C, and absorbance measured after 24, 48, and 72 h of treatment (SynergyLX multi-mode plate reader, Biotek). Each treatment was performed in triplicates. Oxidative stress responses of dosed neurons were determined with Fluorometric Intracellular ROS kit (Sigma-Aldrich #MAK142). Cells were subcultured into Costa-Assay (Corning #3904) clear flat bottom, tissue culture treated, black-coated 96-well plates. Neurons were dosed with active drugs (as described), incubated for 24, 48, or 72 h, and then fluorescence of intracellular ROS was determined following the protocol of the kit using DCFDA as a fluorogenic sensor (lex=650/lem=675nm) (SynergyLX multi-mode plate reader, Biotek). The same protocol was followed using 10 μM Aβ_42_ (Anaspec, #AS24224) in combination with active drugs at 50 μM to quantify intracellular ROS in neurons. Each treatment was performed in triplicates.

##### Imaging

Neurons were analyzed in fluorescence, bright-field, phase contrast, live-cell, and Z-stack imaging with Digital Imaging System Celena-S (Logos). Micrographs were analyzed with Fiji-Image J.

##### Statistical analysis

Data acquired from bioassays were normalized to control for each data set, the normalized average is presented as mean ± standard error of the mean (SEM). SigmaPlot was used for the analysis.

## Data Availability

All study data are included in the article and supporting information. The code is available in GitHub at https://github.com/freshnemo/AD-KG.

## Funding

KH was supported by CPRIT RP140113 (Computational Cancer Biology Training Program Fellowship from Gulf Coast Consortia).

XJ is CPRIT Scholar in Cancer Research (RR180012), and he was supported in part by Christopher Sarofim Family Professorship, UT Stars award, UTHealth startup, the National Institute of Health (NIH) under award number R01AG066749.

YK was supported in part by UTHealth startup and the National Institute of Health (NIH) under award number R01AG066749.

## Author Contributions

XJ and YK provided motivation for the study and contributed to the idea development;

KH, KL, and YK developed the computational method;

KH and KL performed computational experiments;

GPV and GP performed biological experiments;

KH, KL, and YK wrote the initial manuscript;

KH, GPV, KL, GP, XJ, and YK edited the manuscript.

## Conflict of interest

The authors have no conflict of interest to declare.

## SUPPLEMENTARY MATERIALS

Table S1. Ablation study on edge prediction.

Table S2. Node classification task to predict therapeutic targets out of all gene nodes.

Table S3. Drugs with statistically significant treatment effects in reducing AD onset in Optum claim data.

Table S4. Top 100 drugs predicted by multitask learning model and multi-level evidence.

Table S5. Subject’s demographics for population-based treatment effect estimation.

Figure S1. Subgraph of two AD-related genes that were correctly and incorrectly predicted.

Supplementary method 1. Population-based treatment effect

## Notes

### Competing Interest Statement

The authors have declared no competing interest.

### Summary of Updates

Revised the topic, introduction, results, discussion, and methods.

