## Supplementary Tables and Figures for "Synthesize Heterogeneous Biological Knowledge via Representation Learning for Alzheimer’s Disease Drug Repurposing"

**Table S1**. Ablation study on edge prediction. We measured edge prediction accuracy by leaving-one-source-out strategy to quantify the contribution of different interactions.

| Excluded edge type | Predict all edge types using fine-tuned AD knowledge graph | |
|---|---|---|
| | AUROC | AUPRC |
| Drug - target interaction | 0.880 | 0.865 |
| Gene - gene interaction | 0.936 | 0.936 |
| Drug - GO | 0.969 | 0.969 |
| Gene - GO | 0.962 | 0.966 |
| Drug - pathway | 0.679 | 0.728 |
| Drug - drug similarity | 0.981 | 0.979 |
| Gene - pathway | 0.980 | 0.978 |

**Table S2**. Node classification task to predict therapeutic targets out of all gene nodes.

| Therapeutic target node classification | Pretrained universal embedding | AD knowledge graph representation without transfer learning | AD knowledge graph representation with transfer learning |
|---|---|---|---|
| AUROC | 0.642 | 0.937 | 0.947 |
| AUPRC | 0.066 | 0.566 | 0.583 |

**Table S3**. Drugs with statistically significant treatment effects in reducing AD onset in Optum claim data.

| Targets (number of drugs) | Drugs with population-based treatment effect in claim data (ATT) (p-value<0.0001*, p-value<0.001**, p-value<0.05***) |
|---|---|
| Metabolic/vascular risk factors (n=50) | Apixaban (0.1617*), Hydrochlorothiazide (0.1224*), Losartan (0.1175*), Metformin (0.1095*), Salbutamol (0.1025*), Glimepiride (0.0538***) |
| Oxidative Stress (n=13) | Levothyroxine (0.1291*), Atorvastatin (0.1022*) |
| Neuroinflammation (n=13) | Vilanterol (0.1743*), Fluticasone furoate (0.1452*); Methylprednisolone (0.1222*), Montelukast (0.0922**), Ibuprofen (0.0633*), Celecoxib (0.0262*) |
| Neurotransmitters (n=16) | Aminolevulinic acid (0.2088*), Prilocaine (0.1124**), Prochlorperazine (0.1081*), Diazepam (0.094*), Baclofen (0.0857*), Dextromethorphan (0.0820*) |
| Aβ, Tau | Suvorexant (0.1686***), Denosumab (0.1446*), Bordetella pertussis vaccine (0.1398*), hyaluronate sodium (0.1289*), Losartan (0.1175**), Valsartan (0.0579**), Simvastatin (0.0807**), Atorvastatin (0.1021**) |

**Table S4**. Top 100 drugs predicted by multitask learning model and multi-level evidence.

| Rank | Name | Clinical trials | Transcriptomic relevance | Population-based treatment effect | Preclinical trial |
|---|---|---|---|---|---|
| 1 | Galantamine | + | + | | + |
| 2 | Melatonin | + | + | | + |
| 3 | Celecoxib | + | + | | + |
| 4 | Mifepristone | + | | | |
| 5 | Trazodone | + | | + | |
| 6 | Ibuprofen | + | | + | + |
| 7 | Bms 708163 | + | | | |
| 8 | Rosiglitazone | + | + | | + |
| 9 | Amphotericin B | | | | |
| 10 | Physostigmine | | | | + |
| 11 | Minocycline | + | | + | + |
| 12 | Memantine | + | | | + |
| 13 | Indomethacin | + | | | + |
| 14 | Vitamin D | + | | | + |
| 15 | Tacrine | + | | | + |
| 16 | Nevirapine | | | | |
| 17 | Cholinesterase Inhibitors | + | | | + |
| 18 | Phenylephrine | | | | |
| 19 | Acetylcholine | | | | + |
| 20 | Raloxifene Hydrochloride | + | | | |
| 21 | Reserpine | | | | |
| 22 | Icariin | | | | |
| 23 | Nitric Oxide | | | | + |
| 24 | Resveratrol | + | | | + |
| 25 | Heparin, Low-Molecular-Weight | | | | |
| 26 | Curcumin | + | | | + |
| 27 | Norepinephrine | + | | | + |
| 28 | Epinephrine | | | + | |
| 29 | Haloperidol | + | | | |
| 30 | Berberine | | | | + |
| 31 | Entacapone | | | | |
| 32 | Carbachol | | | | |

| | | | | | |
|---|---|---|---|---|---|
| 33 | Streptozocin | | | | |
| 34 | Furosemide | | | + | |
| 35 | Midazolam | + | | | |
| 36 | Maprotiline | | | | |
| 37 | Sumatriptan | | | | |
| 38 | Niacin | | | | |
| 39 | Rivastigmine | + | | | + |
| 40 | Methotrexate | + | | | |
| 41 | Goserelin | | | | |
| 42 | Ceftriaxone | | | + | |
| 43 | Troglitazone | | | | |
| 44 | Ramipril | + | + | | |
| 45 | Glyburide | | | | |
| 46 | Atenolol | | | + | |
| 47 | Sulpiride | | | | |
| 48 | Nicotine | + | | | + |
| 49 | Pioglitazone | + | | | + |
| 50 | Fluorides | | | | |
| 51 | Donepezil | + | | | + |
| 52 | Estradiol | + | | | + |
| 53 | Ticlopidine | | | | |
| 54 | Caffeine | + | | | + |
| 55 | Phenformin | | | | |
| 56 | Dexamethasone | | | + | |
| 57 | Betamethasone | | | + | |
| 58 | Benazepril | | | + | |
| 59 | Carbamazepine | | | | |
| 60 | Thapsigargin | | | | + |
| 61 | Ceramides | | | | |
| 62 | Homocysteine | | | | |
| 63 | Iron | | | | + |
| 64 | Quinine | | | | |
| 65 | Rofecoxib | + | | | + |
| 66 | Cadmium | | | | |
| 67 | Clopidogrel | | | + | |
| 68 | Lovastatin | | | | + |

| | | | | | |
|---|---|---|---|---|---|
| 69 | Ibotenic Acid | | | | |
| 70 | Olanzapine | + | | | |
| 71 | Dopamine | + | | | + |
| 72 | Acetazolamide | | | | |
| 73 | Manganese | | | | + |
| 74 | Sulfasalazine | | | | |
| 75 | Clozapine | | | | |
| 76 | Deferoxamine | | + | | |
| 77 | Pravastatin | + | | + | + |
| 78 | Rosuvastatin Calcium | | | + | |
| 79 | Valdecoxib | | | | |
| 80 | Dipyridamole | | + | | |
| 81 | Progesterone | + | | | + |
| 82 | Etomidate | | + | | |
| 83 | Np 031112 | + | | | |
| 84 | Clonidine | | | | |
| 85 | Hydrochlorothiazide | | | + | |
| 86 | Diglycerides | | | | |
| 87 | Dextromethorphan | + | | | |
| 88 | Trichlorfon | + | | | |
| 89 | Nitrites | | | | + |
| 90 | Triamcinolone | | | + | |
| 91 | Aspartame | | | | |
| 92 | Zinc | | | | + |
| 93 | Ethinyl Estradiol | | | | |
| 94 | Calcitriol | | | | |
| 95 | Baclofen | | | + | |
| 96 | Orphenadrine | | + | | |
| 97 | Terfenadine | | | | |
| 98 | Imipramine | | + | | + |
| 99 | Sodium Salicylate | | | | |
| 100 | 24-Hydroxycholesterol | | | | |

**Table S5.** Subject's demographics for population-based treatment effect estimation. Race, age, and sex distributions before and after matching. Observation ends at AD onset for AD patients.

| Race | | | | | | | | |
|---|---|---|---|---|---|---|---|---|
| | Before Match | | | | After Match | | | |
| | Non-AD | Non-AD % | AD | AD % | Non-AD | Non-AD % | AD | AD % |
| Asian | 44,963 | 3.22% | 4,851 | 2.77% | 4,925 | 2.80% | 4,851 | 2.77% |
| African American | 130,367 | 9.33% | 19,967 | 11.40% | 20,054 | 11.42% | 19,943 | 11.40% |
| Hispanic | 123,579 | 8.84% | 18,862 | 10.77% | 18,949 | 10.79% | 18,830 | 10.76% |
| Unknown | 91,699 | 6.56% | 7,291 | 4.16% | 7,543 | 4.29% | 7,291 | 4.17% |
| Caucasian | 1,006,707 | 72.05% | 124,210 | 70.90% | 124,177 | 70.70% | 124,067 | 70.90% |
| **Gender** | | | | | | | | |
| | Before Match | | | | After Match | | | |
| | Non-AD | Non-AD % | AD | AD % | Non-AD | Non-AD % | AD | AD % |
| Female | 781050 | 55.90% | 107099 | 61.14% | 106941 | 60.88% | 106910 | 61.10% |
| Male | 616085 | 44.09% | 68029 | 38.83% | 68656 | 39.09% | 68020 | 38.87% |
| **Age** | | | | | | | | |
| | Before Match | | | | After Match | | | |
| | Non-AD | Non-AD % | AD | AD % | Non-AD | Non-AD % | AD | AD% |
| 65-69 | 308457 | 22.07% | 6594 | 3.76% | 6661 | 3.79% | 6594 | 3.77% |
| 70-74 | 332466 | 23.79% | 19896 | 11.36% | 19994 | 11.38% | 19896 | 11.37% |
| 75-79 | 315382 | 22.57% | 34805 | 19.87% | 35035 | 19.95% | 34805 | 19.89% |
| 80-84 | 232162 | 16.61% | 57481 | 32.81% | 57224 | 32.58% | 57417 | 32.81% |
| 85-89 | 159833 | 11.44% | 53800 | 30.71% | 53740 | 30.60% | 53665 | 30.67% |
| >=90 | 49015 | 3.51% | 2605 | 1.49% | 2994 | 1.70% | 2605 | 1.49% |
| **Commodities** | | | | | | | | |
| | Before Match | | | | After Match | | | |
| | Non-AD | Non-AD % | AD | AD % | Non-AD | Non-AD % | AD | AD % |
| Head injury | 128910 | 9.23% | 46271 | 26.41% | 46397 | 26.41% | 46111 | 26.35% |
| Heart disease | 139420 | 9.98% | 138631 | 79.14% | 139420 | 79.37% | 138631 | 79.23% |

| | | | | | | | | |
|---|---|---|---|---|---|---|---|---|
| Diabetes | 579364 | 41.46% | 81542 | 46.55% | 81864 | 46.61% | 81439 | 46.54% |
| Vascular disease | 1063184 | 76.09% | 132955 | 75.90% | 134032 | 76.31% | 132955 | 75.98% |
| Obesity | 399385 | 28.58% | 33711 | 19.24% | 33989 | 19.35% | 33711 | 19.27% |
| hypertension | 1192212 | 85.32% | 161653 | 92.28% | 161816 | 92.13% | 161456 | 92.27% |
| hyperlipidemia | 1168458 | 83.62% | 148370 | 84.70% | 148542 | 84.57% | 148185 | 84.69% |

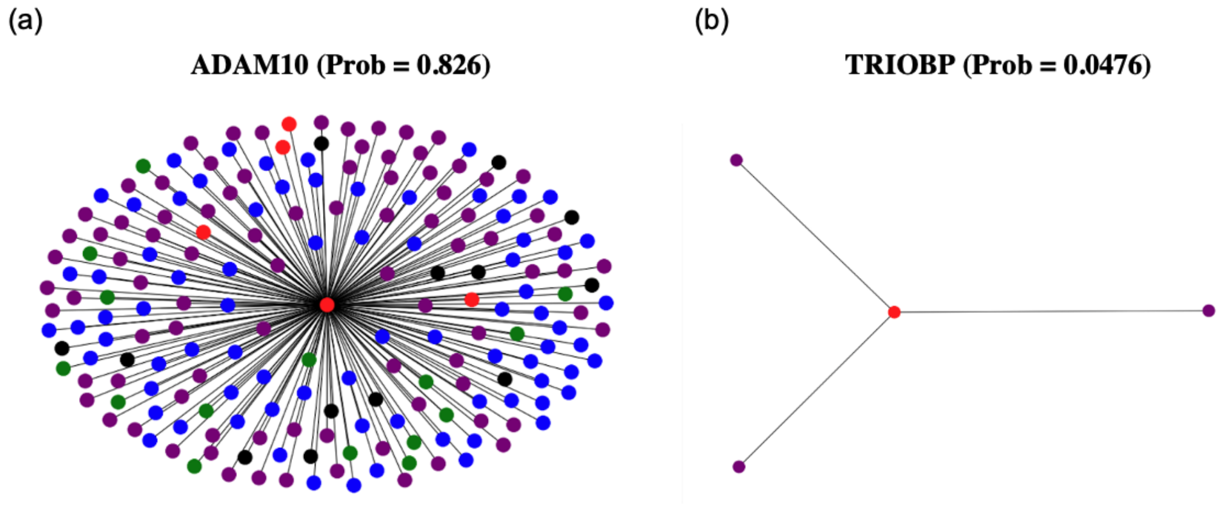


**Figure S1.** Subgraph of two AD-related genes that were correctly and incorrectly predicted. (a) Our model correctly predicted ADAM10 as an AD-related gene. (b) Our model incorrecly predicted TRIOBP as a non-AD gene. (AD-related gene: red; non-AD gene: green; drug: blue; pathway: black; phenotype: purple)

**Supplementary method 1. Population-based treatment effect**

**Database**: We selected our cohort from the Optum Clinformatics® Data Mart subscribed by UTHealth. It comprises administrative health claims from 2007 to mid-2020 for members of a large national managed care company affiliated with Optum. These administrative claims are submitted for payment by providers and pharmacies and are verified, adjudicated, adjusted, and de-identified prior to inclusion in Clinformatics® Data Mart.

**Study subjects.** We included subjects with observation after the age of 65 and observations longer than 5 years. Then we included a total of 350,630 subjects from among AD subjects (n=174,982) and non-AD subjects (n=175,648) that were approximately 1:1 matched, based on age during the observation. Age is a strong risk factor for AD. This window matching is a crucial step to avoid bias caused by age in the censored observation (e.g., short observation does not mean that one is free of complications). That is, for the AD subjects, the observation window started from when any diagnosis code was first recorded and ended when the first AD onset was recorded. For the non-AD subjects, we selected subjects that had the closest observation window by matching age at the observation start and finish. Note that we truncated non-ADRD observations after the age when matched AD observations ended.

**Variables of interest.** Variables of interest were all the comorbid diseases (identified as diagnosis codes) that were diagnosed within the observation window, which might have potential risk to predispose to AD onset. We converted ICD9 or ICD10 diagnosis codes into PheWas codes to increase clinical relevance of the billing codes. PheWas ICD code is a hierarchical grouping of ICD codes based on statistical co-occurrence, code frequency, and human review. For more detail, see reference. We included PheWas disease codes that appeared within the observation window in more than 5% of the subjects. We counted the occurrence of each disease code that appears during the observation and converted them to a logarithm scale (i.e., log2(1+counts)) as the count distributions are skewed. And then we filtered PheWas codes based on the variance of their logarithm of count of occurrence. Finally, a total of 198 PheWas codes were included in the dataset whose occurrence variances are larger than 0.2.

**AD onset.** Outcome of interest was AD onset, which we detected as having either AD diagnosis code or medication. The AD diagnosis codes were PheWas codes for 290.11 (Alzheimer's disease); 290.12 (Frontotemporal dementia, Pick's disease, Senile degeneration of brain); 290.13 (Senile dementia); and 290.16 (Vascular dementia, Vascular dementia with delirium/delusions/depressed mood). Note that we included AD and related dementia because differential diagnosis of these dementias is not correctly recorded in claim data. The AD medications were acetylcholinesterase inhibitors (Donepezil, Galantamine, and Rivastigmine) or memantine.

**Cohort identification.** As a result, of the 68,233,646 patients, there were 896,682 subjects with AD diagnosis codes; 711,006 subjects with AD medication codes; and 1,165,565 subjects with either the diagnosis or medication codes. After excluding subjects without diagnosis/medication codes, timestamp, and observation length less than 5 years, we selected 174,982 AD subjects (case) and matched 175,648 non-AD subjects (control). Cohort matching results in **Table S5** showed that the most common confounders affecting treatment selection (e.g., anti-asthmatic drugs) and AD onset are balanced out.

**Treatment effect estimation.** The counterfactual treatment effect analysis is to identify the difference in outcome (i.e., onset risk) between ones with and without exposure to a certain medication. We denote $Y_i\,(1)$ subject $i$'s outcome when the subject takes the medication ($T_i\ = 1$) and $Y_i\,(0)$ subject $i$'s outcome when the subject does not take the medication ($T_i\ = 0$). The treatment effect $\tau_i$of this medication is defined as: $\tau_i = Y_i\,(1) - Y_i\,(0)$. However, it is impossible to observe factual and counterfactual outcomes at the same time for the same subject. A common approach to mitigate this missing counterfactual outcome is to average out the potential outcomes in the treatment group and control group respectively, by computing propensity scores that represents the probability of treatment assignment based on confounders, and then estimate the average treatment (ATE) effect by $E[Y(1)] - E[Y(0)]$. However, subjects with and without exposure to the medication are not equivalent. We need to balance the differences in covariates between those who received treatment and the comparator so that the likelihood distribution of receiving the treatments is similar between them. We used the inverse probability of treatment weighting (IPTW),[81] which down-weights over-sampled patients and up-weights under-sampled patients so that the treatment group and control group are similar. Specifically, the average treatment effect (ATE) can be estimated through IPTW as

$$\text{ATE}=1/n \sum_{i=1}^{n} \quad [T_i Y_i/e(X_i) - (1-T_i)Y_i/(1-e(X_i)]$$

where $e(X_i)$ is the propensity scores at $T_i = 1$ given subject's features $X_i$. The confounders include demographics and comorbidities (hypertension, diabetes, heart disease, vascular diseases, and brain injury). ATE among the treated or ATT, was calculated the same way but only for patients with $T_i = 1$.

**Drugs with significant treatment effects.** We identified 126 prescribed drugs (out of 530) with positive average treatment effect among treated (ATT) and $p$-value<0.05. We grouped the drugs based on their targets if there is any literature supporting the drug's potential targets.
